# Acute and persistent symptoms in non-hospitalized PCR-confirmed COVID-19 patients

**DOI:** 10.1101/2021.01.22.21249945

**Authors:** Sofie Bliddal, Karina Banasik, Ole Birger Pedersen, Ioanna Nissen, Lisa Cantwell, Michael Schwinn, Morten Tulstrup, David Westergaard, Henrik Ullum, Søren Brunak, Niels Tommerup, Bjarke Feenstra, Frank Geller, Sisse Rye Ostrowski, Kirsten Grønbæk, Claus Henrik Nielsen, Susanne Dam Nielsen, Ulla Feldt-Rasmussen

## Abstract

**Background:** Reports of persistent symptoms after hospitalization with COVID-19 have raised concern of a “long COVID” syndrome. This study aimed at characterizing acute and persistent symptoms in non- hospitalized patients with polymerase chain reaction (PCR) confirmed COVID-19.

**Methods:** Cohort study of 445 non-hospitalized participants identified via the Danish Civil Registration System with a SARS-CoV-2-positive PCR-test and available biobank samples for genetic analyses. Participants received a digital questionnaire on demographics and COVID-19-related symptoms. Persistent symptoms: symptoms >four weeks (in sensitivity analyses >12 weeks).

**Results:** 445 participants were included, of whom 34% were asymptomatic. Most common acute symptoms were fatigue, headache, and sneezing, while fatigue and reduced smell and taste were reported as most severe. Persistent symptoms, most commonly fatigue and memory and concentration difficulties, were reported by 36% of 198 symptomatic participants with follow-up >four weeks. Risk factors for persistent symptoms included female sex (women 44% vs. men 24%, odds ratio 2.7, 95%CI:1.4-5.1, p=0.003) and BMI (odds ratio 1.1, 95%CI:1.0-1.2, p=0.001).

**Conclusion:** Among non-hospitalized PCR-confirmed COVID-19 patients one third were asymptomatic while one third of symptomatic participants had persistent symptoms illustrating the heterogeneity of disease presentation. These findings should be considered in future health care planning and policy making related to COVID-19.

## Introduction

Coronavirus disease 2019 (COVID-19), caused by severe acute respiratory syndrome coronavirus 2 (SARS-CoV-2), has by December 2020 affected more than 82 million people worldwide and has been associated with 2 million deaths leading to a global health and financial crisis [1]. Established risk factors related to dying with the disease include high age, male sex and preexisting comorbidities [2,3,4]. A meta-analysis of 24,410 COVID-19 patients found that the most common symptoms were fever, cough, fatigue, and hyposmia [5]. However, not all exposed individuals catch the disease and not all who have the disease develop symptoms [6,7]. Thus, there is a knowledge gap of individual resilience and risk factors determining the disease trajectory.

In the wake of the initial phase of the COVID-19 pandemic, several observational studies, patient groups and case series have reported persistent symptoms including reduced respiratory capacity [8], fatigue and hyposmia [9,10,11]. In a follow-up study of 143 Italians discharged after hospitalization due to COVID-19, 87.4% still experienced COVID-19-related symptoms at two months after symptom start, and more than half of the patients reported persistent fatigue [12]. Similarly, Jacobs *et al* found that fatigue was the most prevalent symptom 35 days post-hospitalization in 183 participants from the United States, and the majority of participants reported reduced quality of life and both physical and mental health problems [13]. A recent Chinese study presented 6-month follow-up data from 1,733 hospitalized patients, of whom 63% still experienced fatigue and muscle weakness [14]. Clinical and public health interests are therefore no longer limited to information on mortality and clinical outcomes in hospitalized patients, but also to recovery and long-term consequences of COVID-19 post-hospitalization. Among other initiatives, this has led to the Post-Hospitalization COVID-19 study (PHOSP-COVID) in the United Kingdom aiming at long-term follow-up of 10,000 patients discharged after hospitalization due to COVID-19 [15]. The increasing awareness of persistent symptoms among COVID-19 patients has even led to the designation “long COVID” which is yet to be clearly defined [16,17,18]. A recent first draft guideline on long-term effects of COVID-19 published by the National Institute of Health and Care Excellence (NICE) suggests to use the definition “ongoing symptomatic COVID-19” for symptoms lasting between 4 and 12 weeks after the acute onset and “post-COVID-19-syndrome” for symptoms lasting more than 12 weeks [19].

Most studies on COVID-19 symptoms have recruited participants among hospitalized patients or patients attending outpatient clinics or specialized units, or support groups dealing with the consequences of COVID-19 [5,9,20,21]. This confers a selection bias likely overestimating the true prevalence of symptoms and symptom severity. Thus, there is an academic knowledge gap in symptomatology among unselected non-hospitalized patients.

The aim of the present study was to determine the nature and prevalence of acute and persistent symptoms in non-hospitalized patients with COVID-19 confirmed by a positive SARS-CoV-2 PCR.

## Materials and Methods

### Participants

All individuals registered in the Danish Civil Registration System with a COVID-19 diagnosis confirmed by polymerase chain reaction (PCR) for SARS-CoV-2 by August 12^th^ 2020 and an available biobank sample for genetic analyses (n=4,421) were invited via the national digital postbox, e-Boks, to participate in a study on genetics in COVID-19 and related outcomes. The e-Boks is a secure digital postbox used by 92.1% of the Danish adult population by the second quarter of 2020 [22]. A list of SARS-CoV-2 positive individuals was obtained from the Danish Microbiology (MiBa) database, which holds data for all SARS-CoV-2 PCR-tests in Denmark, provided by the Danish Health Data Authority [23,24]. Relevant biobank samples with sufficient material for genetic analyses were identified among stored samples in the Danish National Biobank or the Copenhagen Hospital Biobank [25,26].

Invitations were sent to participants via e-Boks between June 24^th^ and August 15^th^ 2020 together with written project information and a link to a questionnaire on demographic data and symptomatology related to COVID-19. Interested participants could contact a call center between July 27^th^ and August 28^th^ 2020 to receive oral information on the project and provide oral consent prior to receiving a digital link for written consent. The last questionnaire was received digitally by October 31^st^ 2020.

### Inclusion criteria

All individuals in the Danish Civil Registration System with a PCR-confirmed positive test for SARS-CoV-2 and an available biobank sample for genetic analyses with no history of hospitalization due to COVID-19 were eligible for this study (n=4,421). Hospitalization due to COVID-19 was defined as a hospital admission lasting longer than 12 hours within 14 days of a positive PCR-test for SARS-CoV-2. Date of COVID-19 diagnosis and hospitalization status were accessed through the national register for COVID-19 surveillances (EpiMiBa). Non-hospitalized participants with a valid written consent and a completed questionnaire were included in this study.

### Questionnaire

All participants received a link in their e-Boks to a questionnaire with a large number questions on demographic data as well as history and symptoms related to COVID-19. The questionnaire was to a large extent based on a previously validated questionnaire used as part of the Danish Blood Donor Study [27]. Questions on lifestyle factors, demographics and co-morbidities included smoking status, height and weight, occupation, educational level, and the possibility to select the following chronic diseases (using layman terms): Asthma, allergy (other than asthma), diabetes mellitus, hyperthyroidism, hypothyroidism, high blood pressure, heart attack, chest pain (angina pectoris), stroke, chronic bronchitis or big lungs or smokers lungs (emphysema, COPD), osteoarthritis, rheumatoid arthritis, osteoporosis, cancer, other.

Queries on symptomatology related to COVID-19 included the questions “Have you had a feeling of being ill in the period since February 1^st^ 2020?” and if “yes” the following symptoms were listed with answer options of “No”, “Yes, a little”, “Yes, some”, “Yes, a lot”, “Don’t know”: Fever, chills, runny or stuffy nose, reduced sense of smell, reduced sense of taste, sneezing, sore throat, coughing, shortness of breath, headache, muscle and joint pain, chest pain, tiredness and exhaustion [fatigue], difficulties concentrating or remembering, lack of appetite, coloured sputum, red runny eyes, nausea, vomiting, diarrhoea, stomach pain, other. To clarify persistence of symptoms, the questionnaire included the question “Do you still have symptoms” and if the participant answered “yes”, the same list of symptoms was provided albeit without the option of ranking the degree of symptoms.

Symptoms commencing between 28 days before the positive SARS-CoV-2 PCR-test result and 14 days after the test result were included as COVID-19-related symptoms (Supplementary Fig. S1 online). Persistent symptoms were defined as symptoms lasting more than four weeks; these could be ongoing symptoms at the time of filling in the questionnaire in case of a follow-up time of more than four weeks from symptom onset. Alternatively, a stop date could be given in case of a reported symptom lasting more than four weeks from symptom onset to symptom stop. In sensitivity analyses, persistent symptoms lasting more than 12 weeks were explored based on the same selection criteria (12 weeks instead of four weeks). These time limits were in accordance with the time frame for ongoing symptomatic COVID-19 and post-COVID-19-syndrome, respectively, suggested by the National Institute for Health and Care Excellence in the United Kingdom [19].

### Statistics

Demographic data were presented as appropriate by means and standard deviations (SD) or medians and inter-quartile range (IQR). Logistic regression analyses included as covariates: sex (male-female), age, BMI, smoking status (non-smoker vs. ever smoker), comorbidity (yes-no based on composite scores of self-reported chronic disease), work in the health care sector (yes-no), and time to questionnaire (weeks). The latter was included to account for any potential recall bias as the time periods between the PCR test and the questionnaire (in analyses of asymptomatic vs. symptomatic participants) or between the symptom start and the questionnaire (in analyses of persistent vs. no persistent symptoms among symptomatic individuals). Among the health care workers, almost all reported having patient contact [not specified if COVID-related] and thus patient contact was not included in the model. In the logistic regression analyses of risk of acute and persistent symptoms, we used a composite score (any symptom vs no symptom) across all acute and persistent symptoms, respectively. The composite score for acute symptoms included any symptom, regardless of reported severity. P-values reported are based on Welch Two Sample t-test for normally distributed traits and Fisher’s Exact for categorical traits. All statistical analyses and preparation of figures were performed by R version 3.5.0 (the R foundation, www.r-project.org).

### Ethics

The study was part of the “Epidemiology and Genetics of COVID-19” study approved by the National Committee on Health Research Ethics (ID: NVK 2003947) and by the Data Protection Agency (ID: P-2020-356). All participants provided oral and written consent to participate.

## Results

A total of 445 Danish non-hospitalized COVID-19 participants were included (see Fig. 1 for flow chart of the inclusion process).

**Figure 1.**
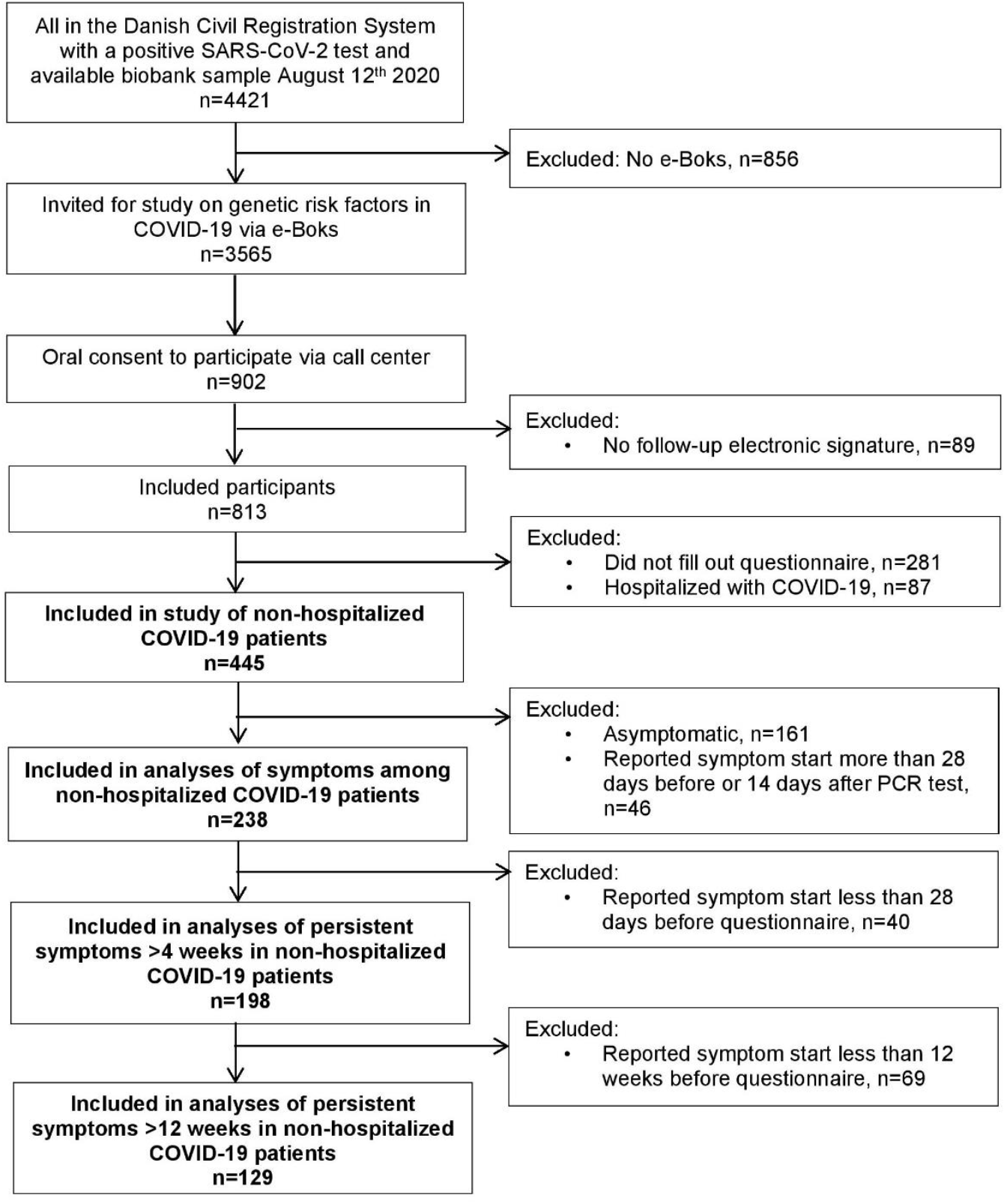
Flowchart of inclusion process. Inclusion process of Danes with PCR-confirmed SARS-CoV-2 in study of symptoms among non-hospitalized patients. **Abbreviations:** e-Boks, secure digital postbox used by 92% of Danes. PCR, polymerase chain reaction. SARS-CoV-2, severe acute respiratory syndrome coronavirus 2.

Most participants had been tested during the Spring of 2020 (Table 1). More women than men participated in the study and significantly more women worked in the health care sector and had patient contact (not specified if COVID-related). Also, the median date for PCR-testing of the included women was one month earlier than that of men (Table 1). There were no other significant differences in demographic data between men and women (Table 1). Less than half of the participants reported preexisting comorbidities; most commonly allergy (28%), hypertension (15%), osteoarthritis (17%), and asthma (8%) (see Supplementary Fig. S2 online).

**Table 1.** Characteristics of participants.

|  | **Women (n=245)** | **Men (n=191)** | **p-value** |
| --- | --- | --- | --- |
| **Age (mean, SD)** | 45.6 (16.1) | 48.7 (16.7) | 0.04 |
| **BMI (median, IQR)** | 24.7 (6.9) | 25.9 (4.6) | 0.42 |
| **Smoking (ever, %)** | 14.2 | 16.2 | 0.59 |
| **Comorbidity (%)** | 55.5 | 56.3 | 1.00 |
| **Work in health care sector (%)** | 37.8 | 7.9 | $5.7 \times 10^{-14}$ |
| **Work with patient contact (%)** | 36.2 | 7.3 | $1.4 \times 10^{-13}$ |
| **Date of positive SARS-CoV-2 test (median, range)** | 22 May 2020<br>(8 Mar 2020 – 12 Aug 2020) | 21 Jun 2020<br>(5 Mar 2020 – 12 Aug 2020) | 0.001 |

Characteristics of included men and women with a history of COVID-19 without need of hospitalization confirmed by a SARS-CoV-2-positive PCR.
Abbreviations: BMI, body mass index. IQR, interquartile range. PCR, polymerase chain reaction. SARS-CoV-2, severe acute respiratory syndrome coronavirus 2. SD, standard deviation.

### Acute symptoms

Completely asymptomatic COVID-19 was reported by 34% of participants. There were no significant differences in sex, smoking status, comorbidities, health care worker status or BMI between those reporting symptoms and those without symptoms. Asymptomatic participants had more often been tested towards the end of the study inclusion period (weeks from PCR to questionnaire; adjusted odds ratio (OR) 1.06 95%CI:1.02-1.09, p=0.0008) (depicted in Supplementary Fig. S3 online), and were slightly older (OR for symptoms 0.98 (0.97-0.99), p=0.004).

Among the 238 participants experiencing symptoms, 55% of women and 52% of men had a sudden onset of symptoms arising within a few hours. Characteristics of symptomatic participants are presented in Table 2. Significantly more symptomatic women than men were working in the health care system, had contact with patients in their work and had been tested slightly earlier in the year (Table 2). The most common symptoms in the acute phase of COVID-19 were fatigue (95%), headache (82%), and sneezing (76%) (Fig. 2A). However, the symptoms most often reported as being severe were reduced smell, fatigue, and reduced taste (Fig. 2A). Excluding all participants with comorbidities did not change the symptom presentation (Fig. 2B).

**Table 2.** Characteristics of non-hospitalized symptomatic men and women with COVID-19.

|  | **Women (n=148)** | **Men (n=90)** | **P-value** |
| --- | --- | --- | --- |
| **Age (mean, SD)** | 44.0 (15.9) | 46.0 (14.8) | 0.32 |
| **BMI (median, IQR)** | 24.3 (6.9) | 25.5 (4.7) | 0.55 |
| **Smoking (ever, %)** | 19.4 | 15.6 | 1 |
| **Comorbidity (%)** | 56.0 | 55.6 | 1 |
| **Work in health care sector (%)** | 44.0 | 13.3 | 8.0x10<sup>-7</sup> |
| **Work with patient contact (%)** | 42.6 | 12.2 | 6.3x10<sup>-7</sup> |
| **Contact to someone with positive test for COVID-19 (%)** | 49.4 | 49.1 | 1 |
| **Symptom start within a few hours (%)** | 55.4 | 52.2 | 0.24 |
| **Date of positive SARS-CoV-2 test (median, range)** | 12-May-2020<br>(10-Mar-2020–12-Aug-2020) | 11-Jun-2020<br>(5-Mar-2020–12-Aug-2020) | 0.14 |
| **Date of symptom onset (median, range)** | 7-May-2020<br>(2-Mar-2020–11-Aug-2020) | 11-Jun-2020<br>(26-Feb-2020–13-Aug-2020) | 0.09 |
| **Time from symptom start to PCR confirmation, days (median, range)** | -2 (-23-6) | -1 (-17-6) | 0.67 |
| **Time from symptom start to questionnaire, days (median, range)** | 104 (5-202) | 69 (7-176) | 0.01 |
| **Time from symptom start to stop, days (median, range) (n=137)** | 14 (1-112) | 12 (1-138) | 0.06 |

Characteristics of 238 non-hospitalized symptomatic men and women with a history of COVID-19 confirmed by a SARS-CoV-2-positive PCR-test.
Abbreviations: BMI, body mass index. IQR, interquartile range. PCR, polymerase chain reaction. SARS-CoV-2, severe acute respiratory syndrome coronavirus 2. SD, standard deviation.

**Figure 2.**
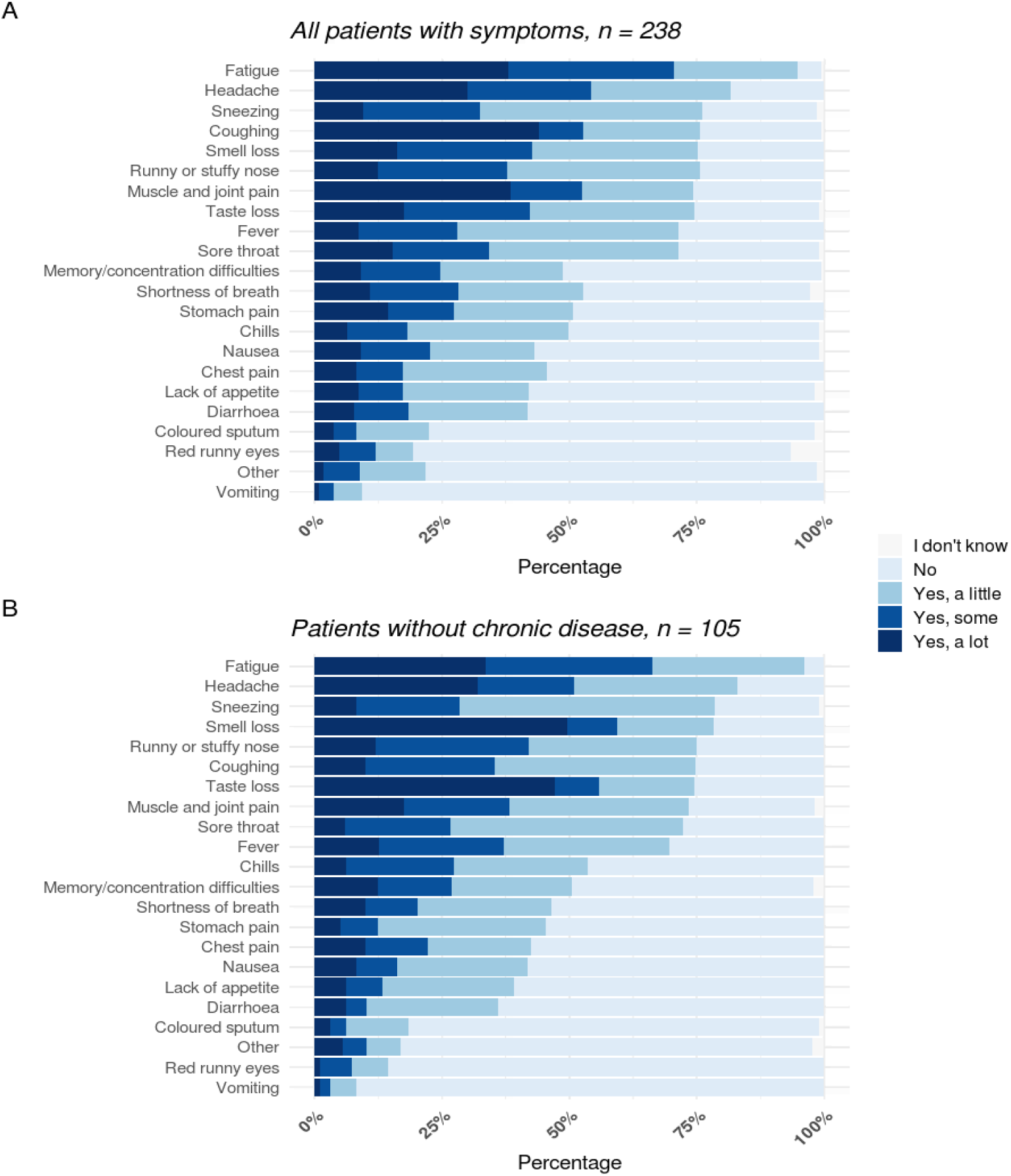
Symptom presentation and severity during acute COVID-19 in non-hospitalized patients. Symptom pattern and severity reported in relation to the acute phase of COVID-19 by non-hospitalized participants with a SARS-CoV-2-positive PCR test. Only participants with symptom start between 28 days before and 14 days after the PCR-test were included. Figure A, all symptomatic participants (n=238). Figure B, symptomatic participants without comorbidities (n=105). Abbreviations: PCR, polymerase chain reaction. SARS-CoV-2, severe acute respiratory syndrome coronavirus 2.

### Persistent symptoms

Most symptomatic participants experienced symptom cessation within two weeks (n=137, women in median 14 days (range:1-112), men in median 12 days (1-138)). Ongoing symptoms at the time of filling in the questionnaire were reported by more than 35% of participants (median time from symptom start 90 days (range 5-197)). Follow-up for more than four weeks was available from 198 symptomatic participants among whom persistent symptoms were reported by 36% (44% of women and 24% of men). The most common persistent symptoms were fatigue (16%), concentration or memory difficulties (13%), reduced sense of smell (10%), and shortness of breath (10%) (Fig. 3). Among participants with more than 12 weeks of follow-up since symptom start (n=129), 40% (48% of women and 23% of men) still had symptoms, especially fatigue (16%) and concentration difficulties (13%), following the same pattern as those with a shorter follow-up period (Fig. 3).

**Figure 3.**
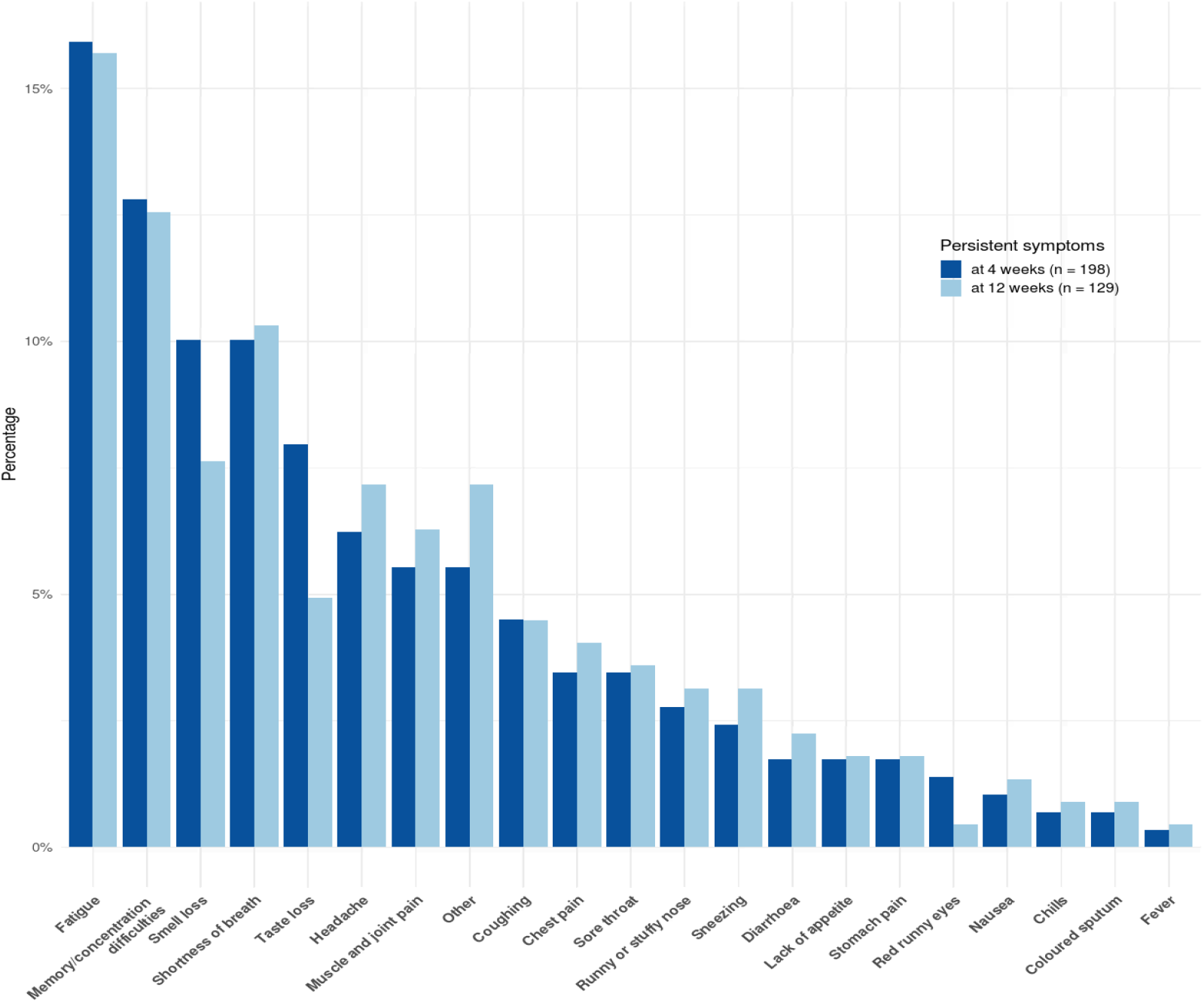
Persistent symptoms in COVID-19 at 4 and 12 weeks. Persistent symptoms after COVID-19. In dark blue, persistent symptoms for more than four weeks in the 198 non-hospitalized participants with symptoms and a follow-up time of more than four weeks and in light blue, persistent symptoms for more than 12 weeks in the 129 non-hospitalized participants with symptoms and a follow-up time of more than 12 weeks.

The risk of persistent symptoms more than four weeks after symptom start was significantly increased in women compared to men (OR 2.7 95%CI:1.4-5.2, p=0.003 (Table 3)). Furthermore, there was an increased risk of persistent symptoms with increasing BMI (OR 1.1 95%CI:1.0-1.2, p=0.001 (Table 3)). No other covariates were significantly associated with the risk of persistent symptoms. Female sex and BMI were also the only significant risk factors for persistent symptoms in the 129 participants with a follow-up period of more than 12 weeks (see supplementary Table S1 available online).

**Table 3.** Risk of persistent symptoms after COVID-19.

|  | **Persistent symptoms (n=72)** | **No persistent symptoms (n=113)** | **Odds ratio (95% CI)** | **p-value** | **Adjusted odds ratio (95% CI)\*** | **p-value** |
| --- | --- | --- | --- | --- | --- | --- |
| **Sex\*\* (women, %)** | 76.4% | 54.9% | 2.66<br>(1.38-5.14) | 0.003 | 2.91<br>(1.32-6.39) | 0.008 |
| **Age (mean (SD))** | 47.4 (15.2) | 45.4 (15.5) | 1.01<br>(0.99-1.03) | 0.38 | 1.00<br>(0.97-1.03) | 0.97 |
| **Smoking (ever, %)\*\*** | 16.7% | 15.0% | 1.13<br>(0.50-2.53) | 0.84 | 0.84<br>(0.30-2.41) | 0.75 |
| **BMI (median (IQR))** | 26.8 (8.6) | 24.4 (4.3) | 1.11<br>(1.04-1.18) | 0.001 | 1.13<br>(1.05-1.22) | 0.001 |
| **Comorbidity (%)** | 47.2% | 42.3% | 1.47<br>(0.77-2.80) | 0.26 | 1.40<br>(0.68-2.85) | 0.36 |
| **Work in health care sector (%)** | 43.1% | 29.5% | 1.81 (0.98-3.36) | 0.08 | 1.28 (0.58-2.81) | 0.54 |
| **Time from symptom start to questionnaire (weeks) (median, range)** | 16<br>(5-25) | 15<br>(5-27) | 1.01<br>(0.96-1.06) | 0.76 | 0.99<br>(0.93-1.06) | 0.86 |

\*The risk of persistent symptoms vs. no persistent symptoms after four weeks in participants with symptoms in the acute phase and a minimum of four weeks of follow-up from symptom start (n=198) adjusted for: sex, age, smoking, BMI, comorbidity and time from symptom start to questionnaire.
\*\*Men as reference group.
\*\*\*Non-smokers as reference group.
Abbreviations: BMI, body mass index. CI, confidence interval. IQR, interquartile range.

More women than men worked in the health care sector (table 1 and 2). Thus, we re-ran analyses of risk factors for persistent symptoms by only including the 117 women with follow-up of more than four weeks. Doing so, BMI was still a significant risk factor (adjusted OR 1.2 95%CI:1.1-1.3, p=0.003), but being a health care worker was not (OR 1.5 95%CI:0.6-3.8, p=0.39).

Furthermore, we depicted acute and persistent symptoms according to sex (Fig. 4). Men and women had a similar prevalence of fatigue in the acute phase (Fig. 4 left hand side). In participants with persistent symptoms, fatigue was the most common persistent symptom in both men and women, but the prevalence of persistent fatigue was almost twice as high in women compared to men (28% vs. 15%, OR 2.2 95%CI:1.0-4.7, p=0.05, right hand side of Fig. 4).

**Figure 4.**
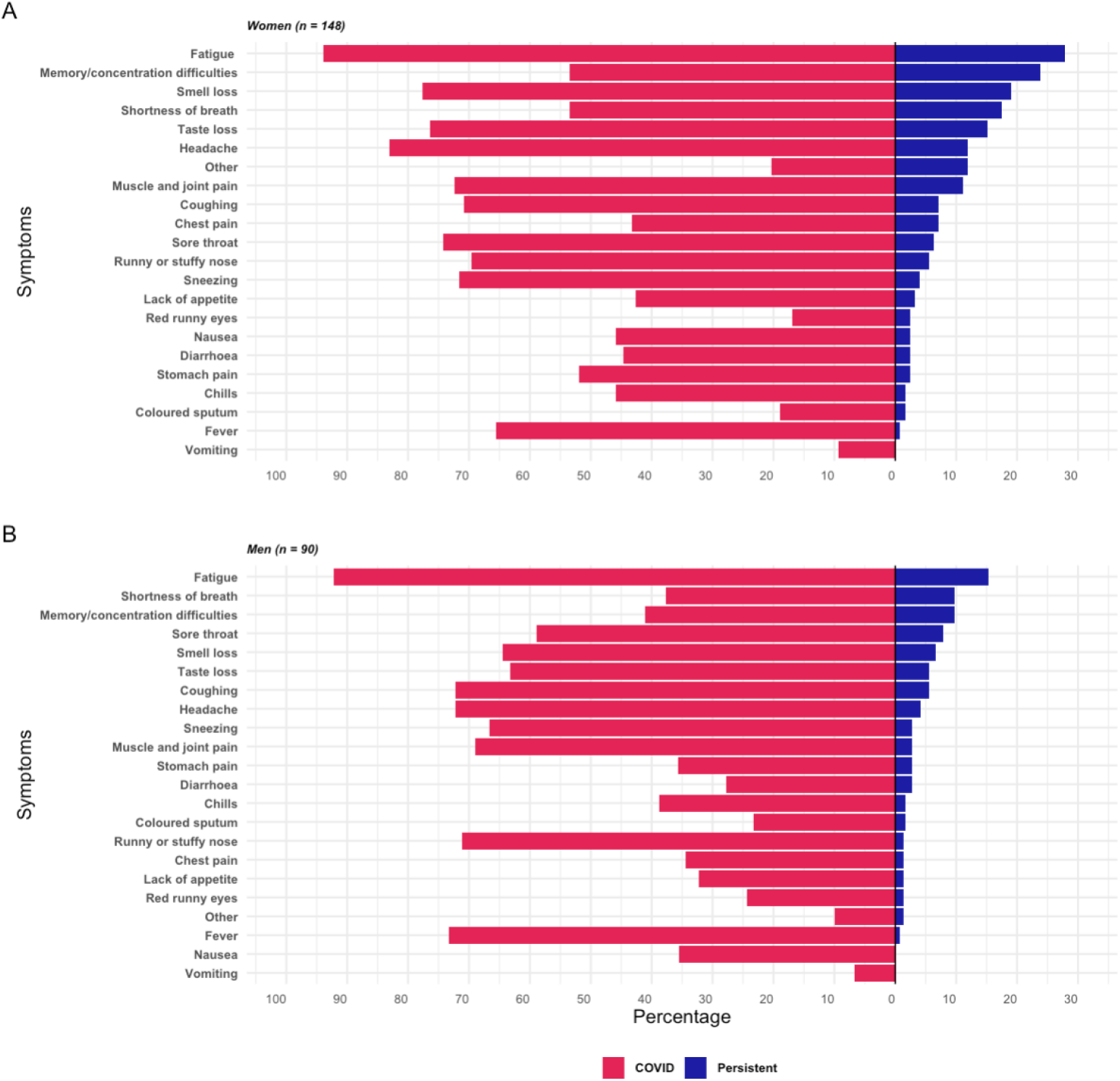
Acute and persistent symptoms in non-hospitalized men and women with COVID-19. The percentage of acute (red panel to the left) and persistent (blue panel to the right) symptoms in non-hospitalized COVID-19 participants according to sex. Persistent symptoms were defined as lasting more than four weeks. A, prevalence of reported acute (total n=148) and persistent symptoms (total n=126) in women. B, prevalence of reported acute (total n=90) and persistent symptoms (total n=72) in men.

## Discussion

The present study demonstrated a high prevalence of persistent symptoms in non-hospitalized patients with COVID-19 confirmed by SARS-CoV-2 PCR-testing. The most common persistent symptoms were fatigue and memory and concentration difficulties with a higher risk in women. Furthermore, there was an equally high prevalence of asymptomatic COVID-19 participants.

Unlike most previous studies on COVID-19-related symptoms an important strength of the present study was the recruitment of non-hospitalized participants with a positive SARS-CoV-2 PCR-test. Thus, we were able to depict variations in symptoms even among non-hospitalized participants with a confirmed COVID-19 diagnosis. Contrary to studies relying solely on information gathered through questionnaire data, our use of national register data on SARS-CoV-2 PCR tests and hospitalization strengthens the validity of the study results. However, our study finding of women being at higher risk of persistent symptoms may be biased by the fact that more asymptomatic individuals, especially men, were included towards the end of the inclusion period and were slightly older. This likely reflects both an altered test strategy in Denmark with easier access to targeted testing for mild cases of COVID-19 and screening of close contacts, and a survival bias with a higher risk of hospitalization among older citizens and symptomatic men. In addition, more women worked in the health care sector with patient contact and although this was not associated with an increased risk of persistent symptoms in the present study, larger studies are needed to clarify if the increased risk of persistent symptoms among women could be confounded by specific lines of work. Finally, all studies of COVID-19- related symptoms suffer from an inherent likelihood of attracting participants with a personal experience of severe or persistent symptoms thus risking an overestimation of the true prevalence of persistent symptoms. A similar selection bias may be present in our study, however, likely reduced compared to most studies specifically addressing COVID-19-related symptoms because we specifically invited participants to a study on genetics and not symptoms.

Our finding of fatigue as the most prevalent persistent symptom matches previous studies among post-hospitalized COVID-19 patients suffering from fatigue [9,12,28,29]. To our knowledge, few studies have been published on persistent symptoms in non-hospitalized patients with positive SARS-CoV-2 PCR-tests. Among 292 participants interviewed by phone at a median of 16 days post-testing, Tenforde *et al* found 35% of participants with acute symptoms to still be experiencing symptoms (especially coughing, fatigue and shortness of breath) [30]. In a study of 180 participants (8 of whom had been hospitalized) interviewed during the acute phase of disease and at follow-up phone calls, persistent symptoms were reported by 53% of symptomatic participants at a mean follow-up time of 125 days (most often fatigue, loss of smell and taste, and arthralgias) [31]. Larger survey-based studies on COVID-19-related symptoms in a multitude of participants with self-reported COVID-19 are available online, although not yet peer-reviewed (Table 4 presents an overview of studies) [32,33,34,35,36,37]. Characteristic for most of these studies is the acceptance of self-reported COVID-19 status by participants without laboratory confirmation of COVID-19-status, and recruitment of participants with the specific aim of COVID-19 symptom reporting likely inferred a substantial selection bias. However, most studies do report fatigue, concentration problems and loss of smell and taste as the most common persistent symptoms, which is comparable to our findings in PCR-confirmed COVID-19 non-hospitalized participants. Thus, our study provides substantial laboratory- and registry-confirmed evidence in support of these symptoms being part of a “long COVID”-syndrome existing even in non-hospitalized COVID-19 patients.

**Table 4.** Overview of studies on persistent symptoms in non-hospitalized COVID-19.

| Author | Participants* | Recruitment | Diagnosis of COVID-19 | Persistent symptoms |  |  |  |  |
| --- | --- | --- | --- | --- | --- | --- | --- | --- |
|  |  |  |  | Reporting | Definition | Prevalence | Most common | Risk factors |
| Bliddal (present study) | 445 non-hospitalized | PCR-positive cases invited to genetics study | Positive PCR confirmed by national register | Questionnaire at time of inclusion | 4 weeks<br>12 weeks | 36%<br>40% | Fatigue, memory and concentration difficulties, smell loss, shortness of breath | Female, BMI |
| Cirulli (32) ** | 233 (8 hospitalized) (3,652 negative controls, 17,474 non-tested) | Population study initiated before COVID-pandemic | Self-reported positive COVID-test | Prospective survey every 4-6 weeks | 30 days<br>60 days<br>90 days | 42.3%<br>33.8%<br>24.1% | Anosmia, ageusia, memory loss, and headache | Severity of acute phase, initial dyspnoea |
| Davis (33)** | 3,762 (56.7% non-hospitalized) with illness >28 days | Social media and support groups for long COVID | Suspected or confirmed COVID-19 cases (27% laboratory-confirmed) | Survey distribution via online COVID-19 support groups and social media | 28 days<br>90 days<br>6 months | 100%<br>96%<br>65.2% | Fatigue, post-exertional malaise, and cognitive dysfunction | 78.9% female responders |
| Goërtz (36) | 2,113 (2,001 non-hospitalized) | Facebook groups for persistent coronavirus and website of Lung Foundation | Confirmed (n=345), symptom-based or suspected COVID-19 | Online questionnaire link available in recruitment fora | 3 weeks (mean 79±17 days) | 99.3% | Fatigue, dyspnoea | Age, number of acute symptoms, previous health status, comorbidities |
| Patient Research Group (35) ** | 640 (56.4% no hospital contact) | Online survey on long COVID | Tested for COVID-19 (n=324) or attributing symptoms to COVID-19 | Online survey on symptoms and demographics related to COVID-19 | Mean ongoing 40 days, recovery 27 days | NA | Shortness of breath, tightness of chest, fatigue, brain fog/concentration challenges (most prevalent at week 4) | 76.6% female responders |
| Petersen (31) | 180 (8 hospitalized) | PCR-positive cases invited to long COVID study | Positive PCR | Phone interview(s) at inclusion and follow-up (symptom onset to last interview: mean days 125 (SD 18), range:45-215) | Mean 125 days | 53.1% (8.9% severe persistent) | Fatigue, loss of smell and taste, and arthralgias | Number of acute symptoms, age 50-66 years |
| Sudre (34)** | 4,182 incident cases from UK, USA, and Sweden (% hospitalization NA) | Population symptom monitoring during COVID-pandemic | Self-reported positive PCR | Prospective weekly symptom logging in app | 4 weeks<br>8 weeks<br>12 weeks | 13.3%<br>4.5%<br>2.3% | Fatigue, headache, dyspnoea and anosmia | Age, BMI, female, hospital assessment |
| Tenforde (30) | 292 outpatients | Random sample of PCR-positive outpatients | Positive PCR at outpatient clinic | Telephone interview 14-21 days after PCR-test | Median 16 (14-21) days | 35% of symptomatic participants | Cough, fatigue, congestion, dyspnoea | Age, psychiatric condition, BMI, comorbidities |
| Walsh-Messinger (37)** | 52 students at private university (96 non-COVID students) | Students invited for online survey | Suspected or confirmed (75% PCR-positive) | Online surveys at inclusion | ≥28 days | 51% | Impaired concentration, headache, rhinitis, exercise intolerance, dyspnoea, sleep impairment, brain fog, appetite loss, fatigue, and chest pain | Female, acute symptom severity |

\*Not included: Studies of outpatients recruited via COVID-units or specialized clinics due to COVID-19-related symptoms (i.e. smell loss) or follow-up, case reports or series, studies without hospitalization or symptom status, non-English language. \*\*Study available online included due to relevance despite unpublished non-peer-reviewed. Abbreviations: NA, not available. PCR, polymerase chain reaction for severe acute respiratory syndrome coronavirus 2.

Why SARS-CoV-2 carries a high risk of persistent symptoms, especially in women, is an urgent research question. Suggested mechanisms fall into two categories of either direct effects from the virus or indirect effects caused by immune responses to the virus [38]. While persistent symptoms including chronic fatigue are well-described in other pulmonary infections such as SARS and pneumonia, SARS-CoV-2 may differ in its multi-organ-affection and high contagiousness [38,39,40,41]. The female preponderance bears resemblance to that of many autoimmune diseases [42]. Unfortunately, the present study did not have laboratory data to assess either initial or persistent immunological response against SARS-CoV-2 and did not have data for retesting for SARS-CoV-2 to ascertain resolution of the viral load in participants with persistent symptoms. However, previous studies have found a poor correlation between the risk of persistent symptoms and objective measures of disease despite performing a wide range of clinical and laboratory tests in COVID-19 patients either during the acute phase or at follow-up [9,28]. Also, a negative SARS-CoV-2 PCR-test as a sign of recovery has been criticized for not corresponding to the patients’ perception of (or lack of) recovery [33,35]. Thus, detailed laboratory data and functional studies are urgently needed to understand the immunological component of long COVID.

An important question arising from the present and other studies of long COVID is how to manage these patients with persistent symptoms. Previous studies of chronic fatigue have shown uncertain benefit of treatment such as exercise, cognitive behavioral therapy, and rehabilitation initiatives, and are, nevertheless, not necessarily compatible with post-COVID fatigue [43,44,45]. Several specific management initiatives are emerging for “long COVID” as well as guideline initiatives by the National Institute for Health and Care Excellence in United Kingdom, and guidance material for self-rehabilitation after COVID-19 released by the World Health Organization [9,12,19,46]. Some have argued for the importance of involving general practitioners in the management of this syndrome [47] and some have advocated for the importance of including patients in emerging new strategies [16,48]. A subset of patients has yet to recover after months of symptoms and those patients may need long-term follow-up at specialized units. Thus, an important aspect of future studies on this matter is to identify patients at risk of long-term symptoms. The present study suggests that up to 40% of non-hospitalized previously healthy patients will experience some degree of persistent symptoms with an increased risk in women. Larger studies of big data including real-time symptom reporting will aid in determining risk factors and predicting disease trajectories [34,49]. The mounting evidence of “long COVID” calls for active policy making to secure prevention and early detection, facilitate optimal treatment and support rehabilitation [16,38].

Equally important may be the high proportion of asymptomatic COVID-19 individuals. In the present study 34% of non-hospitalized PCR-confirmed SARS-CoV-2-positive participants reported no symptoms in the acute phase of the disease. However, the overall prevalence of asymptomatic COVID-19 was similar to that estimated in Danish health care workers (assessed by IgM and IgG antibodies against SARS-CoV-2) of 46.5% [50]. Similarly, a study of the Icelandic population found that 43% were asymptomatic at time of a positive SARS-CoV-2 PCR test among participants included in a population screening unlike targeted testing of severely symptomatic individuals [51]. Furthermore, the COVID Symptom Score study prospectively monitored more than 2 million citizens in UK, US and Sweden. In a subset of 26,495 participants with a self-reported positive SARS-CoV-2 PCR-test, 55.3% were asymptomatic [34,49] [numbers deduced from supplementary files]. Although some studies reported a lower prevalence [31], there did seem to be a general agreement on a true high prevalence of asymptomatic COVID-19. In a recent analytical model based on the assumption that 30% of COVID-patients may by asymptomatic, Johansson *et al* found that asymptomatic individuals would carry 50% of all transmissions [52]. This has important implications for disease control and stresses the need for active tracing and testing of close contacts as part of disease control.

In conclusion, even among non-hospitalized PCR-confirmed SARS-CoV-2-positive participants, more than one third reported persistent symptoms after four and 12 weeks, respectively. The most prevalent persistent symptoms were fatigue and memory and concentration difficulties. Contrary to this, 34% of non-hospitalized patients were asymptomatic in the acute phase of the disease stressing the importance of testing near contacts. The high prevalence of both patients with persistent symptoms and of asymptomatic patients bear witness to the heterogeneity of the disease presentation. Although data should be reproduced due to possible limitations of survival or recall bias, these findings should be taken into account in future health care planning and policy making related to COVID-19 prevention, detection, treatment and follow-up.

## Data Availability

The data that support the findings of this study are available from The Danish COVID-19 Genetic Consortium but restrictions apply to the availability of data used under license for the current study, and thus are not publicly available. Data are however available from the authors upon reasonable request and with permission from The Danish COVID-19 Genetic Consortium under Danish legislation.

## Acknowledgements

We highly appreciate our collaboration with the Danish COVID-19 Genetic Consortium (Supplementary Appendix 1 online). Work related to the present study was supported by a grant from Sygesikring Danmark (2020-0178) to the Danish COVID-19 Genetic Consortium and by the Independent Research Fund Denmark (0216-00014B to NT). KB, LC, DW, MS and SB (Brunak) acknowledge the Novo Nordisk Foundation (grants NNF17OC0027594 and NNF14CC0001). KG was supported by a grant from the European Hematology Association. SDN received research grants from Novo Nordic Foundation, FSS, Lundbeck Foundation, and Rigshospitalet Research Council. UFR’s research salary was supported by a research grant from The Kirsten and Freddy Johansen’s Fund. The funding sources had no impact on the planning or conduction of the study or of any work related to this manuscript.

## Author contributions

SB and KB planned and conducted the analyses and drafted the manuscript and revisions. IN, HU, SDN, OBP, SB and MT contributed to the participant inclusion. OBP, SDN, and UFR developed the questionnaire. MS was in charge of project data management. All coauthors contributed to the conception or design of the work, or the acquisition, analysis, or interpretation of data, as well as reviewed and approved the content of the manuscript.

## Additional Information

The authors declare no competing interests.

## References

1. Johns Hopkins University. The Johns Hopkins Coronavirus Resource Center. https://coronavirus.hu.edu/map.html. (2020).

2. Wu, Z. & McGoogan, J. M. Characteristics of and Important Lessons From the Coronavirus Disease 2019 (COVID-19) Outbreak in China: Summary of a Report of 72314 Cases From the Chinese Center for Disease Control and Prevention. JAMA. 323, 1239–1242 (2020).

3. Huang, C. et al. Clinical features of patients infected with 2019 novel coronavirus in Wuhan, China. Lancet. 395, 497–506 (2020).

4. Zheng, Z. et al. Risk factors of critical & mortal COVID-19 cases: A systematic literature review and meta-analysis. J. Infect. 81, e16–e25 (2020).

5. Grant, M. C. et al. The prevalence of symptoms in 24,410 adults infected by the novel coronavirus (SARS-CoV-2; COVID-19): A systematic review and meta-analysis of 148 studies from 9 countries. PLoS. One. 15, e0234765 (2020).

6. Kronbichler, A. et al. Asymptomatic patients as a source of COVID-19 infections: A systematic review and meta-analysis. Int. J. Infect. Dis. 98, 180–186 (2020).

7. Arons, M. M. et al. Presymptomatic SARS-CoV-2 Infections and Transmission in a Skilled Nursing Facility. N. Engl. J. Med. 382, 2081–2090 (2020).

8. Fraser, E. Long term respiratory complications of covid-19. BMJ. 370, m3001 (2020).

9. Townsend, L. et al. Persistent fatigue following SARS-CoV-2 infection is common and independent of severity of initial infection. PLoS. One. 15, e0240784 (2020).

10. Hopkins, C. et al. Six month follow-up of self-reported loss of smell during the COVID-19 pandemic. Rhinology. doi: 10.4193/Rhin20.544. Online ahead of print (2020).

11. Fjaeldstad, A. W. Prolonged complaints of chemosensory loss after COVID-19. Dan. Med. J. 67 (2020).

12. Carfi, A., Bernabei, R., & Landi, F. Persistent Symptoms in Patients After Acute COVID-19. JAMA. 324, 603–605 (2020).

13. Jacobs, L. G. et al. Persistence of symptoms and quality of life at 35 days after hospitalization for COVID-19 infection. PLoS. One. 15, e0243882 (2020).

14. Huang, C. et al. 6-month consequences of COVID-19 in patients discharged from hospital: a cohort study. Lancet. Published Online January 8, 2021, 1-12 (2021).

15. PHOSP-COVID consortium & Leicester University. The Post-hospitalisation COVID-19 study (PHOSP-COVID). https://www.phosp.org/. (2020).

16. The Lancet. Facing up to long COVID. Lancet. 396, 1861 (2020).

17. BMJ. Long covid: How to define it and how to manage it. https://www.bmj.com/content/370/bmj.m3489. doi: https://doi.org/10.1136/bmj.m3489 (2020).

18. Nature. Long COVID: let patients help define long-lasting COVID symptoms. Nature. 586, 170 (2020).

19. National Institute for Health and Care Excellence. COVID-19 rapid guideline: managing the long-term effects of COVID-19. https://www.nice.org.uk/guidance/ng188/chapter/1-Identifying-people-with-ongoing-symptomatic-COVID-19-or-post-COVID-19-syndrome. 2020).

20. Meys, R. et al. Generic and Respiratory-Specific Quality of Life in Non-Hospitalized Patients with COVID-19. J. Clin. Med. 9, 3993 (2020).

21. Ladds, E. et al. Persistent symptoms after Covid-19: qualitative study of 114 “long Covid” patients and draft quality principles for services. BMC. Health Serv. Res. 20, 1144 (2020).

22. The Danish Digitization Agency. Statistics on Digital Post. https://digst.dk/it-loesninger/digital-post/om-loesningen/tal-og-statistik-om-digital-post/. (2020).

23. Voldstedlund, M., Haarh, M., & Molbak, K. The Danish Microbiology Database (MiBa) 2010 to 2013. Euro. Surveill. 19, 20667 (2014).

24. Statens Serum Institut. the Danish Microbiology Database. https://miba.ssi.dk/service/english. (2021).

25. Statens Serum Institut. The Danish National Biobank. https://www.danishnationalbiobank.com/. (2020).

26. The Capital Region of Denmark. The Copenhagen Hospital Biobank. https://www.regionh.dk/forskning-og-innovation/Region-Hovedstadens-biobank/Sider/default.aspx. (2020).

27. Pedersen, O. B. et al. SARS-CoV-2 infection fatality rate among elderly retired Danish blood donors - A cross-sectional study. Clin. Infect. Dis. Online ahead of print (2020).

28. van den Borst, B. et al. Comprehensive health assessment three months after recovery from acute COVID-19. Clin. Infect. Dis. Online ahead of print (2020).

29. Mandal, S. et al. ‘Long-COVID’: a cross-sectional study of persisting symptoms, biomarker and imaging abnormalities following hospitalisation for COVID-19. Thorax. Online ahead of print (2020).

30. Tenforde, M. W. et al. Symptom Duration and Risk Factors for Delayed Return to Usual Health Among Outpatients with COVID-19 in a Multistate Health Care Systems Network - United States, March-June 2020. MMWR Morb. Mortal. Wkly. Rep. 69, 993–998 (2020).

31. Petersen, M. S. et al. Long COVID in the Faroe Islands - a longitudinal study among non-hospitalized patients. Clin. Infect. Dis. Online ahead of print (2020).

32. Cirulli, E. T. et al. Long-term COVID-19 symptoms in a large unselected population. https://www.medrxiv.org/content/10.1101/2020.10.07.20208702v3. (2020).

33. Davis, H. E. et al. Characterizing Long COVID in an International Cohort: 7 Months of Symptoms and Their Impact. https://www.medrxiv.org/content/10.1101/2020.12.24.20248802v2. (2020).

34. Sudre, C. H. et al. Attributes and predictors of Long-COVID: analysis of COVID cases and their symptoms collected by the Covid Symptoms Study App. https://www.medrxiv.org/content/10.1101/2020.10.19.20214494v2. (2020).

35. Patient-Led Research for COVID-19. Report: What Does COVID-19 Recovery Actually Look Like? https://patientresearchcovid19.com/research/report-1/. (2021).

36. Goertz, Y. M. J. et al. Persistent symptoms 3 months after a SARS-CoV-2 infection: the post-COVID-19 syndrome? ERJ. Open. Res. 6, doi.org/10.1183/23120541.00542-2020 (2020).

37. Walsh-Messinger, J. et al. The Kids Are Not Alright: A Preliminary Report of Post-COVID Syndrome in University Students. https://www.medrxiv.org/content/10.1101/2020.11.24.20238261v2.full.pdf. (2020).

38. Nature Medicine. Meeting the challenge of long COVID. Nat. Med. 26, 1803 (2020).

39. Metlay, J. P. et al. Measuring symptomatic and functional recovery in patients with community-acquired pneumonia. J. Gen. Intern. Med. 12, 423–430 (1997).

40. Lam, M. H. et al. Mental morbidities and chronic fatigue in severe acute respiratory syndrome survivors: long-term follow-up. Arch. Intern. Med. 169, 2142–2147 (2009).

41. Tansey, C. M. et al. One-year outcomes and health care utilization in survivors of severe acute respiratory syndrome. Arch. Intern. Med. 167, 1312–1320 (2007).

42. Billi, A. C., Kahlenberg, J. M., & Gudjonsson, J. E. Sex bias in autoimmunity. Curr. Opin. Rheumatol. 31, 53–61 (2019).

43. Turner-Stokes, L. & Wade, D. T. Updated NICE guidance on chronic fatigue syndrome. BMJ. 371, m4774 (2020).

44. Larun, L., Brurberg, K. G., Odgaard-Jensen, J., & Price, J. R. Exercise therapy for chronic fatigue syndrome. Cochrane. Database. Syst. Rev. 10, CD003200 (2019).

45. Vink, M. & Vink-Niese, A. Could Cognitive Behavioural Therapy Be an Effective Treatment for Long COVID and Post COVID-19 Fatigue Syndrome? Lessons from the Qure Study for Q-Fever Fatigue Syndrome. Healthcare. (Basel). 8, 552 (2020).

46. World Health Organization. Support for Rehabilitation: Self-Management after COVID-19 Related Illness. https://www.who.int/publications/m/item/support-for-rehabilitation-self-management-after-covid-19-related-illness. (2020).

47. Greenhalgh, T., Knight, M., A’Court, C., Buxton, M., & Husain, L. Management of post-acute covid-19 in primary care. BMJ. 370, m3026 (2020).

48. Gorna, R. et al. Long COVID guidelines need to reflect lived experience. Lancet. (2020).

49. Drew, D. A. et al. Rapid implementation of mobile technology for real-time epidemiology of COVID-19. Science. 368, 1362–1367 (2020).

50. Iversen, K. et al. Risk of COVID-19 in health-care workers in Denmark: an observational cohort study. Lancet Infect. Dis. 20, 1401–1408 (2020).

51. Gudbjartsson, D. F. et al. Spread of SARS-CoV-2 in the Icelandic Population. N. Engl. J. Med. 382, 2302–2315 (2020).

52. Johansson, M. A. et al. SARS-CoV-2 Transmission From People Without COVID-19 Symptoms. JAMA Netw. Open. 4, e2035057 (2021).

